# Development of a novel musculoskeletal hypothesis using sparse Group Factor Analysis: the ADVANCE cohort

**DOI:** 10.1101/2025.07.02.25330713

**Authors:** Fraje CE Watson, Fabio S Ferreira, Balasundaram Kadirvelu, Alex N Bennett, Aldo A Faisal, Neil Graham, Harriet Kemp, Paul Cullinan, Christopher Boos, Nicola T Fear, Anthony MJ Bull

## Abstract

Musculoskeletal conditions are a leading global cause of disability, yet the factors influencing long-term musculoskeletal health, particularly following trauma, remain incompletely understood. This study applies sparse Group Factor Analysis, a hierarchical unsupervised machine learning method, to the ADVANCE cohort—a longitudinal dataset of 1445 UK Afghanistan War servicemen—to identify latent structures in multimodal clinical data. Study 1 validated the approach by rediscovering known group-level patterns between combat-injured and non-injured participants, including poorer outcomes in pain, mobility, and bone health among those with lower limb loss. Study 2 explored the Injured, non-amputee subgroup without prespecified labels to identify new hypothesis-generating clusters that could subsequently be tested using standard hypothesis testing methods. A subgroup of 125 individuals with worse musculoskeletal outcomes was uncovered. This group had greater body mass, higher injury severity, and a higher prevalence of head injury. These findings led to a novel hypothesis: that head injury, including potential traumatic brain injury, is associated with long-term musculoskeletal deterioration. This hypothesis is supported by literature in both athletic and military populations and will be tested in follow-up analyses. Our findings demonstrate how sparse Group Factor Analysis, combined with clinical insight, can uncover hidden patterns in large-scale datasets and generate testable, clinically relevant hypotheses that inform prevention, treatment, and rehabilitation strategies.

**Author Summary:** Musculoskeletal conditions such as osteoarthritis and low back pain are the second largest contributor to global disability. They can be caused by a variety of factors such as ageing, genetics, lifestyle, and injury. Understanding the interconnectedness of long-term musculoskeletal outcomes following injury could help improve prevention, intervention and rehabilitation initiatives to reduce resulting disability. In this study, we describe a new machine learning methodology called Sparse Group Factor Analysis that we apply to a complex dataset from a military cohort study to generate new research hypotheses. The first study (n=1145) validated our approach by generating hypotheses that we had already investigated via traditional methods. The second study used a sub-set of the cohort (125 participants with poor musculoskeletal outcomes). This showed a link between poor musculoskeletal outcomes and head injury, resulting in a new hypothesis that a head injury or traumatic brain injury may contribute to poor musculoskeletal outcomes. We will test this hypothesis using traditional methods in follow-up analyses. We have demonstrated how Spare Group Factor Analysis can be used alongside clinical knowledge to find hidden patterns in in large, complex datasets to provide information that could inform improved prevention of future musculoskeletal injury, intervention and rehabilitation strategies.

## Introduction

Musculoskeletal health is an important but complex societal issue. Musculoskeletal disorders are associated with ageing (1), genetics (2), occupation (3, 4), trauma (5, 6), and lifestyle (7), and can be exacerbated by psychosocial factors (8). Musculoskeletal injury can further compound these challenges (5). Components associated with increased risk of musculoskeletal injury are multifactorial, but include: physical fitness, obesity and being underweight (9), age, sex, (10) sports such as running (10) and contact sports (11), ergonomics (12), poor neuromuscular control (13), and physically demanding occupations (e.g., military) (14, 15).

Musculoskeletal conditions are the second-highest cause of global non-fatal disability (16, 17). Musculoskeletal conditions affect more than a third of the UK population (18) and 494 million people globally (16). They result in 2,462 disability-adjusted life years per 100,000 globally, which is increasing over time (16, 19, 20).

Better understanding of the interconnectedness of long-term musculoskeletal outcomes (e.g., two outcomes influence each other, one outcome is associated with a particular injury type) associated with injury could guide preventative, interventional and rehabilitation efforts, potentially decreasing the disability burden and increase quality life years. Achieving this better understanding would require large, longitudinal datasets. However, looking beyond the expected to generate novel associations or hypotheses in a large dataset is challenging. Artificial Intelligence (AI) could provide the necessary processing powering and pattern recognition skills required to overcome this. AI has already shown diagnostic, treatment personalisation and prognostic potential in medicine across a number of diagnoses (e.g., detecting breast cancer, personalising sepsis treatment strategies, predicting progression of neurodegenerative diseases) (21–29).

Unsupervised machine learning, a branch of AI, focuses on uncovering hidden patterns and intrinsic structures within unlabelled datasets, without relying on labelled data (30). Key techniques within unsupervised learning include clustering, dimensionality reduction, and density estimation. These methods are particularly valuable in exploratory data analysis as they enhance model interpretability and efficiency by elucidating latent structures and reducing complexity. Here, we use sparse group factor analysis (GFA), a hierarchical probabilistic machine learning model to identify subgroup-specific and subgroup-common multimodal latent disease factors in participant groups (31).

The ADVANCE study is collecting prospective, longitudinal physical and psychosocial data from a cohort of 1,145 UK servicemen who served in the Afghanistan War (32) with the aim of reporting the long-term physical and psychosocial outcomes of combat-related injury. The baseline dataset of ADVANCE contains over 13 million datapoints. Important, predetermined hypotheses that set the recruitment target numbers for the study have been made and tested using musculoskeletal data (6, 33–35). The application of sparse GFA unsupervised machine learning techniques have the potential to uncover hidden patterns in complex datasets to generate previously unseen hypotheses, thus addressing one of the key limitations of analytical techniques currently in use.

This paper will describe the methodology undertaken to identify a novel and clinically relevant hypothesis using the musculoskeletal data of the ADVANCE study. To do this, we present two studies: (1) application of sparse GFA to the ADVANCE dataset and comparison of results to known data patterns, and (2) application of sparse GFA to a nested study using a sub-group of heterogenous and under-investigated ADVANCE participants to generate a novel hypothesis. We hypothesise that GFA can be applied to a complex musculoskeletal dataset and that new clinically relevant hypotheses on causation/correlation can be inferred de novo.

## Results

### Demographic information

Across the entire cohort, mean age was 34.1-years (SD: 5.4-years), mean height was 179.1cm (SD: 6.8cm), mean (adjusted) body mass was 89.0kg (SD: 13.3kg). Participants self-described their race as White (n=1036, 90.6%), Black (n= 46, 4.0%), Asian (n= 41, 3.6%), or Mixed/Other (n= 21, 1.8%). Data were collected from Injured participants (those who sustained combat injuries in the Afghanistan war requiring aeromedical evacuation to the UK) participants a mean of 8.3-years (SD: 2.1-years) post-injury and from Uninjured participants (frequency matched participants who did not sustain combat injuries in the Afghanistan war requiring aeromedical evacuation to the UK) a mean of 7.7-years (SD: 1.9-years) since matched deployment.

### Sparse Group Factor Analysis

#### Study 1

Fourteen data components were identified (Supplementary Material A), of which the top four are shown in Figure 1. Table 1 shows the dominant variables that were different between Injured and Uninjured groups within each factor and overall interpretation.

**Figure 1:**
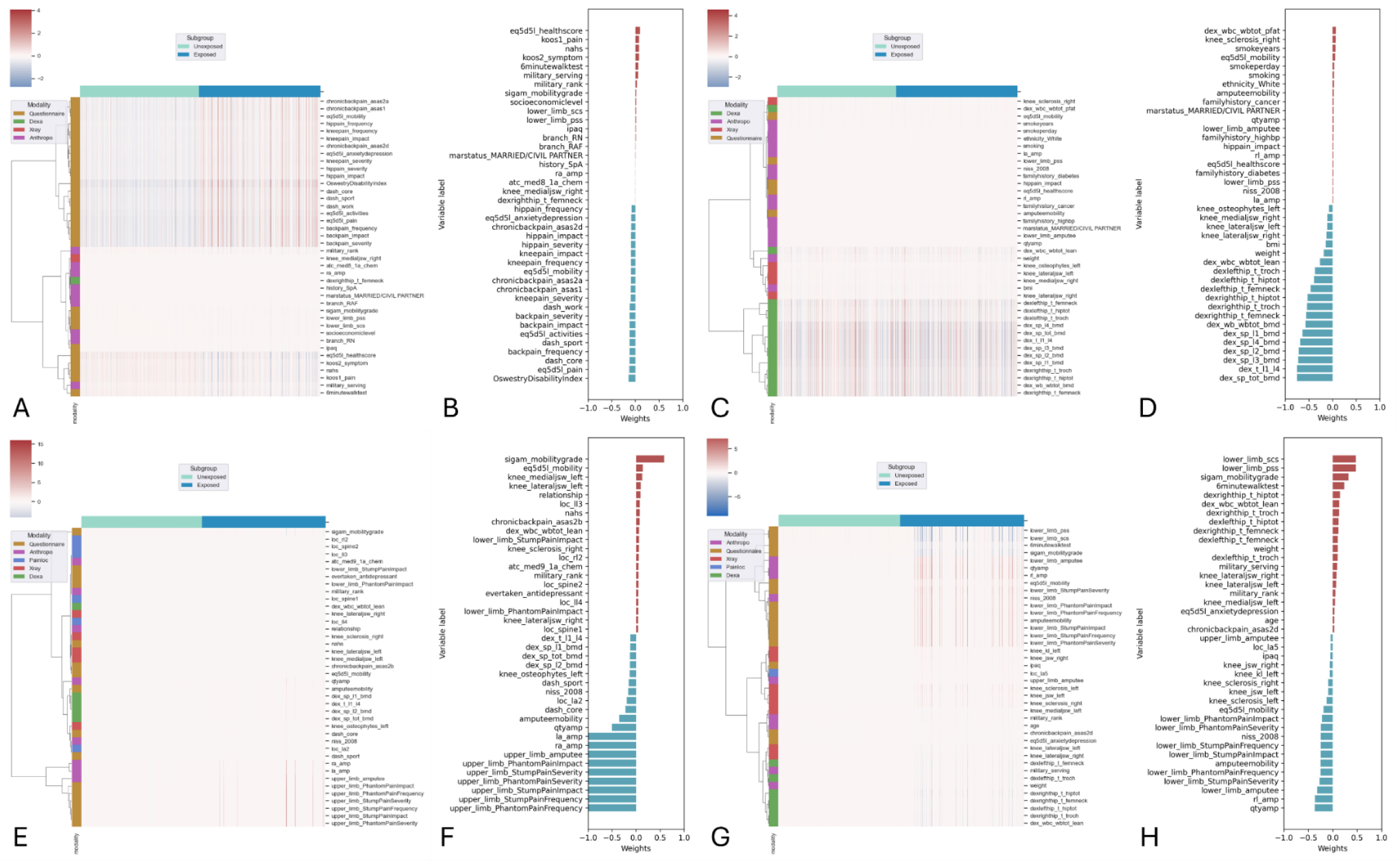
Study 1 - Top four data components identified by sparse Group Factor Analysis in the Uninjured and Injured participants in the ADVANCE dataset and their associated weights (A & B, C & D, E & F, G & H).

**Table 1:**
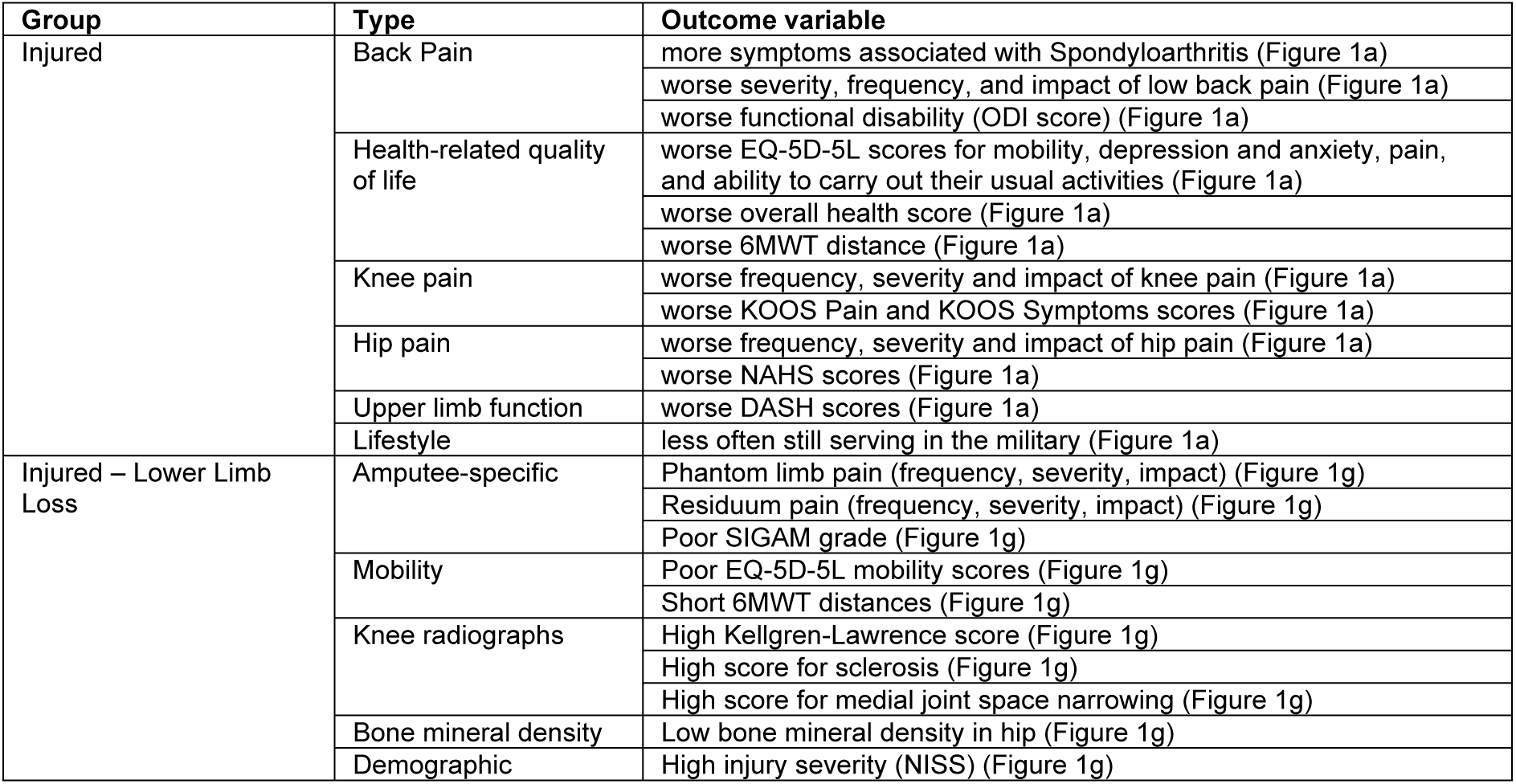
Data components identified in Study 1 analysis, which can be seen in the raw data output in Figure 1.

In Figure 1c and Figure 1d, bone mineral density was identified as a key component, but it was not explained by a difference between Injured and Uninjured groups.

Many of these clusters have already been investigated using hypothesis-driven statistical analyses by the ADVANCE study. For example, we have previously reported higher knee and hip Kellgren-Lawrence, DASH, ODI, overall pain, and back pain scores in the Injured group (6, 34, 36–38); and that participants with lower limb loss have significantly lower bone mineral density in their amputation-side femoral neck (33).

#### Study 2

Fourteen data components were identified (Supplementary Material B), of which the top four are shown in Figure 2.

**Figure 2:**
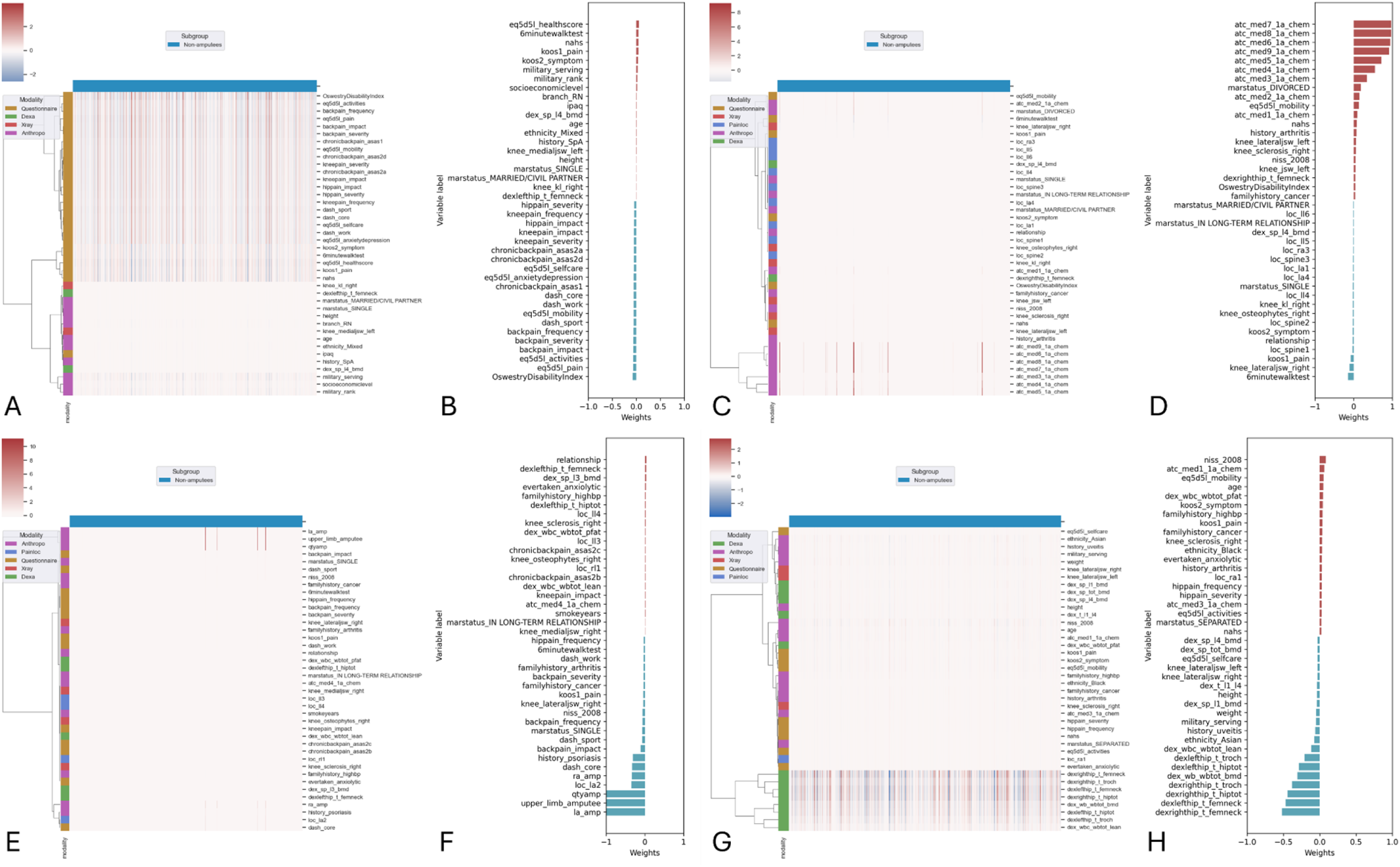
Study 2 - Top four data components identified by sparse Group Factor Analysis in the Injured, non-amputee participants in the ADVANCE dataset and their associated weights (A & B, C & D, E & F, G & H).

Following the steps described above, a group structure was identified in Figure 2a, which identified a group with high (red) scores across multiple outcomes and low (blue) scores for some other outcomes (Table 2). Next, participants were split into two groups along the identified structure. In this instance, this led to a group of 125 participants with poor musculoskeletal outcomes, and the remaining 297 without. Finally, patterns in the demographic, injury pattern and overall health outcome data were used to identify possible reasons for worse musculoskeletal outcomes.

**Table 2:** Data components identified in Round 2 analysis.

| Group | Type | Outcome variable |
| --- | --- | --- |
| Sub-group of 125 | Back Pain | more symptoms associated with Spondyloarthritis (Figure 2a) |
|  |  | worse severity, frequency, and impact of low back pain (Figure 2a) |
|  |  | worse functional disability (Figure 2a) |
|  | Health-related quality of life | worse EQ-5D-5L scores for mobility, depression and anxiety, pain, and ability to care for themselves (Figure 2a) |
|  |  | worse overall health score (Figure 2a) |
|  |  | worse 6MWT distance (Figure 2a) |
|  | Knee pain | worse frequency, severity and impact of knee pain (Figure 2a) |
|  |  | worse KOOS Pain and KOOS Symptoms scores (Figure 2a) |
|  | Hip pain | worse frequency, severity and impact of hip pain (Figure 2a) |
|  |  | worse NAHS scores (Figure 2a) |
|  | Upper limb function | worse DASH scores (Figure 2a) |
|  | Demographic | Less often still serving in the military (Figure 2a) |
|  |  | Lower military rank (Figure 2a) |

##### Pattern identification

There were no significant differences in age (mean: 34.5-years vs. 34.2-years), height (mean: 179.1cm vs. 178.6cm), or time since injury (8.6-years vs. 8.6-years) between groups with and without poor musculoskeletal outcomes. The group with poor musculoskeletal outcomes were heavier (mean: 92.6kg, SD: 14.7kg) and had higher NISS scores (median: 12, IQR: 5-22) than those without (mean weight: 88.0kg, SD: 13.4kg; median NISS: 9, IQR: 4-14), both *p*=0.002.

Descriptively, the group with poor musculoskeletal outcomes contained a higher percentage of participants with upper limb (56.0% vs. 42.7%), abdominal (28.44% vs. 16.9%), head (18.4% vs. 12.9%) and spinal injuries (21.1% vs. 12.1%) compared to the group with better musculoskeletal outcomes. For injury combination, both groups had the same top three: lower extremity injuries only (13.8% and 24.6%), a combination of upper and lower extremity injuries (7.3% and 7.4%), and upper extremity injuries only (6.4% and 7.0%) (Figure 3).

**Figure 3:**
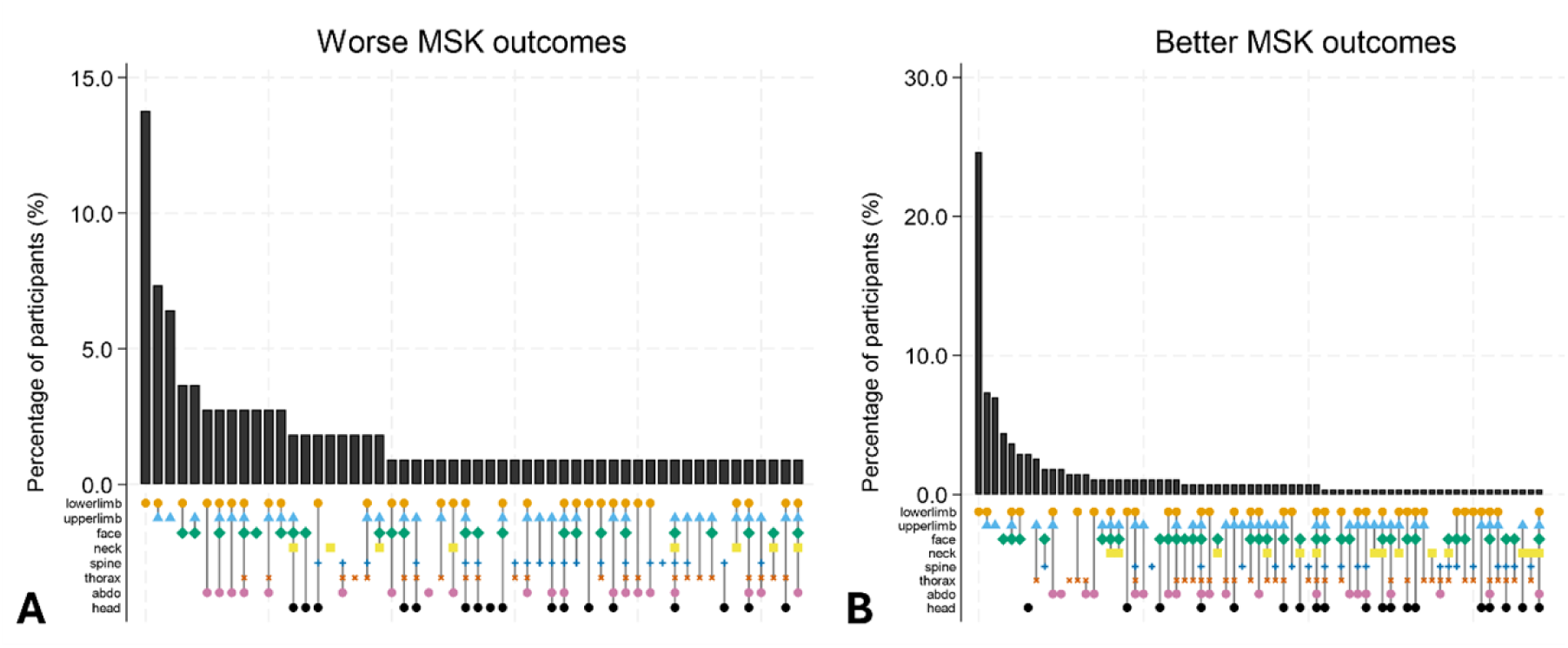
Upset plot showing injury location combinations for participants in the Injured, non-amputee cohort with (A) and without (B) poor musculoskeletal outcomes.

Visual inspection of boxplots further split each group by presence or absence of an injury in each location and compared overall health score. As expected, scores for those with poor musculoskeletal outcomes were always worse, however, head injury (Figure 4d) was the only body region to worsen the overall health score further in the group with poor musculoskeletal outcomes. Full results in Supplementary File B).

**Figure 4:**
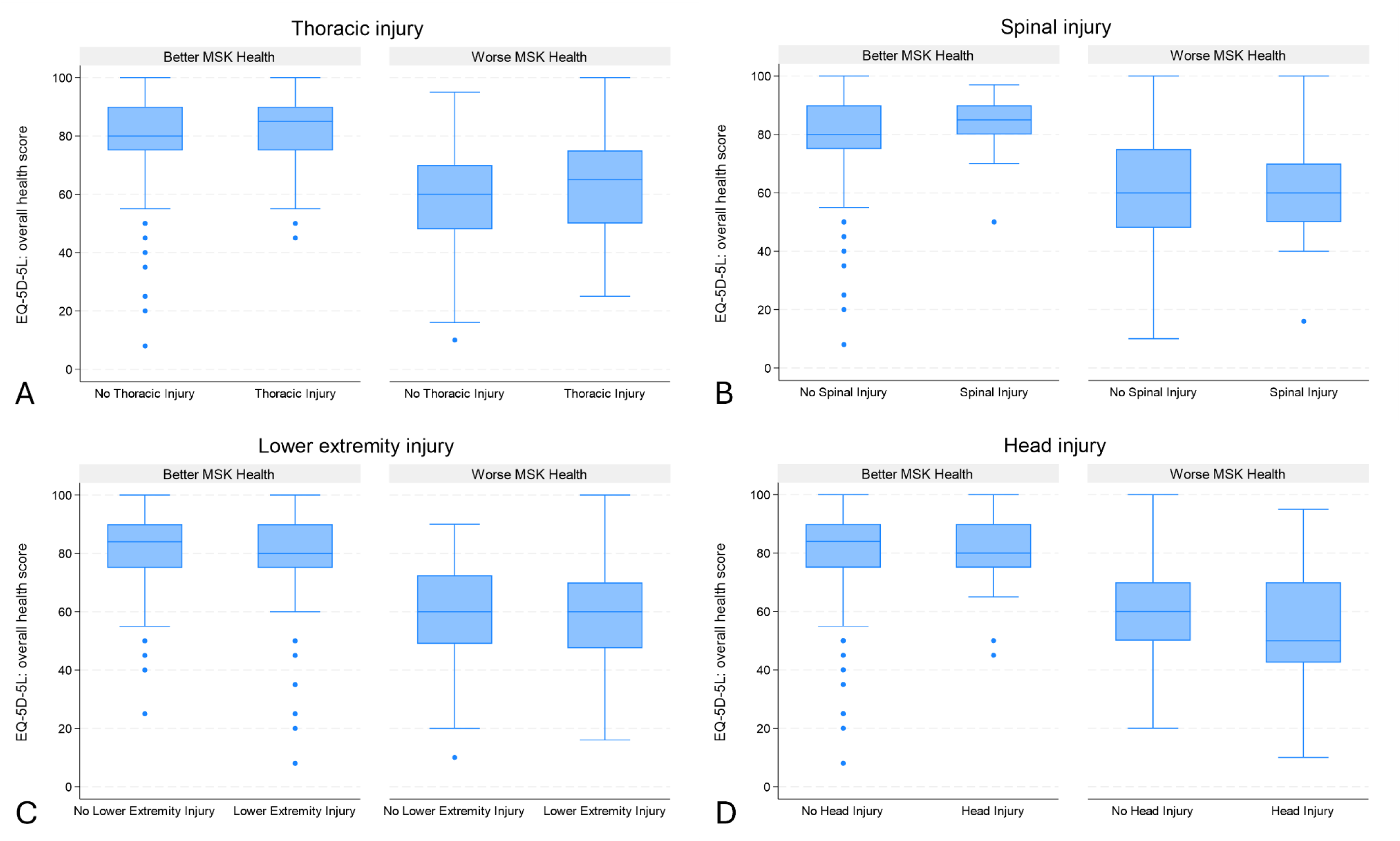
Boxplot plot of overall health score for Injured, non-amputee participants with and without poor musculoskeletal outcomes, by presence/absence of a thoracic (A), spinal (B), lower extremity (C) or head (D) injury.

Pattern identification revealed the following hypotheses regarding worse musculoskeletal outcomes in a sub-group of the Injured – non-amputees: greater body mass, worse injury severity, and head injury.

## Discussion

This work has demonstrated a method for generating novel hypotheses in multi-dimensional data using sparse GFA. In Study 1, we showed how sparse GFA was able to identify known clusters in the cohort, thus corroborating their value. Study 2 showed how sparse GFA was able to identify a cluster with particularly poor musculoskeletal outcomes in a cohort with a heterogenous internal structure. We investigated patterns in the cluster to generate hypotheses about the potential cause. Specifically, in a cluster of participants with worse musculoskeletal outcomes, we identified higher injury severity score, higher body mass, and head injury as potential causes. The novel head injury hypothesis could be investigated further using typical statistical methods.

In Study 1 using the full ADVANCE cohort, we were able to identify patterns that had already been addressed, thus confirming the validity and utility of the model. Many of these findings were heavily influenced by scores from the participants with lower limb loss, consistent with the ADVANCE study’s research where their musculoskeletal outcomes are often worse (6, 33–38). Sparse GFA also generated hypotheses on hip pain across the Injured and Uninjured group that were already being investigated when this study was performed. Study 1 identified a pattern in bone mineral density findings that was not explained by Injured/Uninjured status. Future consideration of that pattern, such as that demonstrated in Study 2, could be applied here to formulate a hypothesis.

Having confirmed the validity of sparse GFA, we subsequently analysed the Injured-, non-amputee group separately as they are a more heterogenous group without a clear internal structure that has not been investigated. Using one loading matrix as an example, sparse GFA identified a group of 125 participants with similarly poor musculoskeletal outcomes. Exploration of patterns delineating these groups found that those with worse musculoskeletal outcomes were significantly heavier, which is logical as obesity is often linked to worse musculoskeletal outcomes (39). The group with worse musculoskeletal outcomes had significantly worse injury severity scores, which also explains worse outcomes. Using injury and overall health outcome data, we identified head injury as a possible cause of worse musculoskeletal outcomes. This methodology has identified a new hypothesis for the ADVANCE study; long-term musculoskeletal outcomes of participants who sustained a head injury are worse than participants who did not.

A head injury could cause a traumatic brain injury either as a result of direct impact to the head or secondary to blast (40). A review of the literature on head injuries, traumatic brain injury or concussion and musculoskeletal injury established that this is a developing field in professional athletes and with existing reports on military personnel. A systematic review and meta-analysis found a two-times greater odds of musculoskeletal injury in athletes following a concussion up to two years later (41), with similar findings in a meta-analysis of papers including athletes and military personnel (42). Military personnel who sustained a concussion (i.e. a mild traumatic brain injury) were 1.84-times more likely to sustain an upper limb musculoskeletal injury in the subsequent year (43). A long-term outcome study in people with a moderate-to-severe traumatic brain injury >15-years prior report high rates of musculoskeletal complaints (44). There is no consensus on the neuropathology of these findings, but some studies suggest impaired neuromuscular control (45) and difficulty with dual-task control (46) following traumatic brain injury.

Based on the findings from this study, the association between head or facial injury, traumatic brain injury, and poor musculoskeletal outcomes in military personnel will be investigated in the ADVANCE cohort using hypothesis-driven statistical methods. The density of prospective, longitudinal musculoskeletal outcome data with longer follow-up times than most existing literature will ensure a unique contribution to the literature. These findings will inform guidance on the treatment of traumatic brain injury, long-term outcomes, and identification of areas for rehabilitation.

Sparse GFA could be applied to a larger, more complex dataset across multiple healthcare-related modalities such as mental health, cardiovascular health, proteomics, and MRI brain imaging. Many of the clusters and hypotheses formed in this pilot study had already been addressed because it is likely that different musculoskeletal outcomes will be related to each other. Introducing more data types across specialities could uncover a previously unseen or unknown interconnectedness between data themes. This method blends the high processing power of AI whilst retaining a rational clinical input to generate novel hypotheses that can be further tested using traditional statistical techniques.

### Limitations

This study identified injury locations using the JTTR, which has limitations as the information is documented in a live war zone and active trauma care setting, which could result in inaccuracies. There is potential that injuries are missed and not documented at this stage, such as head injuries. Additionally, other injuries could contribute to traumatic brain injury, not just head injury. Future testing of this new hypothesis will incorporate a thorough and robust analysis of potential traumatic brain injury to more accurately define head injury status.

The JTTR defines head and face injuries separately, so only head injuries contributed to this hypothesis, but some facial injuries are also linked to traumatic brain injury (47). The ADVANCE study is currently working on defining traumatic brain injury using information from the JTTR, self-reported past medical history, and the Ohio State Questionnaire, with severity defined using the Mayo Classification System. This classification will allow for a comprehensive assessment of traumatic brain injury and thorough investigation of the hypothesis.

The overall health today outcome of the EQ-5D-5L was used to test the effect of injury location as it was not body region specific. Doubts have been cast on the EQ-5D-5L, which has been considered to have a ceiling effect and to be insensitive to change in some populations, despite its strong psychometric properties (48). Finally, the use of ADVANCE musculoskeletal data only could have biased the results compared to using the full ADVANCE dataset, but this is inherent to the pilot nature of this work and we plan to apply it to the full dataset in time.

### Conclusion

This application of sparse GFA combined with human interpretation can be used to identify hidden patterns and intrinsic structures within complex, multi-modality datasets to generate hypotheses that may have not been evident previously. We have demonstrated this method with a pilot portion of musculoskeletal data from the ADVANCE study and generated a novel hypothesis regarding head injury.

## Materials & Methods

### Ethics Statement

Ethical approval was granted by the UK Ministry of Defence Research Ethics Committee (357/PPE/12). Informed, written consent was gained for each participant.

### The ADVANCE cohort

The protocol for the ADVANCE study is described elsewhere (32). Participants (n=1145) were male serving or veterans of the UK military deployed on combat operations to Afghanistan between 2003 and 2014. Injured participants (n=579, 50.6%) sustained combat injuries requiring aeromedical evacuation to the UK. Uninjured participants (566, 49.4%) were frequency matched by deployment, service, rank, role and age to Injured participants to form a comparison group. The study is limited to men, since fewer than 18 female personnel sustained combat injuries, of whom only three had serious or very serious injuries (49).

Baseline data were collected between March 2016 and August 2020, a median of approximately 8-years post-injury or matched deployment.

Comprehensive physical and psychosocial data were collected from all participants across a range of domains such as cardiovascular, mental health, respiratory health, musculoskeletal health, proteomics, hearing, and ageing. This pilot analysis uses the musculoskeletal-adjacent data described in Table 3.

**Table 3:** Type, name, and description of ADVANCE variables used to assess ADVANCE study participants and entered into the sparse Group Factor Analysis model.

| Method | Type | Details |
| --- | --- | --- |
| Clinical assessment and self-reported | Demographic | Age, height, (adjusted) body mass, New Injury Severity Score (NISS), socioeconomic status, body mass index, abdominal circumference, waist to hip ratio, race, service, role, rank, smoking status, amputation status |
| Dual x-ray absorptiometry (DEXA) of the whole body, right and left proximal femur, and lumbar spine | Bone mineral density | t-score for the left and right hip (femoral neck, trochanter, ward's triangle, intertrochanter, hip total), t-score for the L1-L4 spine, raw bone mineral density for each lumbar vertebrae (L1-L4), bone mineral density for the whole body |
|  | Body composition | Total lean tissue in the whole body, percentage of fat tissue in the whole body |
| Posterior-anterior semi-flexed knee radiographs | Radiographic knee osteoarthritis | Kellgren-Lawrence score (0-4)(52), Osteoarthritis Research Society International (OARSI) joint space narrowing score (0-3), OARSI osteophyte score (0-3), OARSI sclerosis score (0-3)(53), minimum medial joint space width, and minimum lateral joint space width for left and right knees |
| Physical assessment – all participants | Six-minute walk test | Furthest distance a participant can walk in 6-minutes |
| Physical assessment – lower limb loss only | Amputee Mobility Predictor (54) | Ability to perform 21 tasks scored by independent assessor scored 0-47 where a higher score is indicative of better mobility |
| Questionnaire – all participants | Assessment of SpondyloArthritis International Society (ASAS) definition of inflammatory back pain (55) | 5 questions about back pain to identify inflammatory back pain (suspected when $\geq 4$ answers are yes) |
|  | Health-related quality of life score (EQ-5D-5L) (56) | Health-related quality of life score for individual items; mobility, self-care, usual activities, pain/discomfort, anxiety/depression each scored 1-5 where a higher score is a worse outcome, and overall health scored 0-100 where 100 is the best health. |
|  | Back pain | Scored for severity, frequency and impact 0-10 where 10 is the severe/frequent/impactful |
|  | Oswestry Disability Index (ODI)(57) | 10-item questionnaire each scored 0-5 with a total score of 0-100 where 100 is indicative of the most back pain related disability. |
|  | Knee Osteoarthritis Outcome Score (KOOS) – Pain (58) | 9-item questionnaire each scored 0-5 for left and right knees, each with a total score of 0-100 where 0 is indicative of the worst pain. |
|  | Knee Osteoarthritis Outcome Score (KOOS) – Symptoms (58) | 7-item questionnaire each scored 0-5 for left and right knees, each with a total score of 0-100 where 0 is indicative of the worst symptoms. |
|  | Knee and Hip Joint Pain scores | Left and right knees and hips scored for severity, frequency and impact 0-10 where 10 is the severe/frequent/impactful |
|  | Non-arthritic Hip Score (NAHS) | 20-item questionnaire each scored 0-5 for left and right hips, each with a total score of 0-100 for each hip where 0 is indicative of the worst hip pain/symptoms/activities/function. |
|  | Disability of the Arm, Shoulder and Hand (DASH) – Core (59) | 30-item questionnaire each scored 0-5 with a total score of 0-100 where 100 is indicative of the most disability |
|  | Disability of the Arm, Shoulder and Hand (DASH) – Work (59) | 4-item questionnaire each scored 0-5 with a total score of 0-100 where 100 is indicative of the most disability |
|  | Disability of the Arm, Shoulder and Hand (DASH) – Sport (59) | 4-item questionnaire each scored 0-5 with a total score of 0-100 where 100 is indicative of the most disability |
|  | International Physical Activity Questionnaire (IPAQ) (60) | Total weekly activities categorised as moderate (“activity that makes you breathe somewhat harder than before”) and vigorous (“activities that take hard physical effort and make you breathe much harder than normal”) |
| Questionnaire – lower limb loss specific | Special Interest Group in Amputee Medicine (SIGAM) Mobility Questionnaire (61) | Scored A-F based on self-reported mobility |
|  | Socket comfort score (62) | Scored per amputated limb 0-10 where 10 is the most comfortable |
|  | Prosthetic satisfaction score (62) | Scored per amputated limb 0-10 where 10 is the most satisfaction |
|  | Phantom pain score | Scored for severity, frequency and impact 0-10 where 10 is the severe/frequent/impactful |
|  | Stump pain score | Scored for severity, frequency and impact 0-10 where 10 is the severe/frequent/impactful |

For participants with limb loss, adjusted body mass was calculated depending on body segments missing (50, 51) and without wearing prosthetics. The height of participants with lower limb loss was measured using actual or reported height.

Injuries sustained by the Injured group were identified using the Joint Theatre Trauma Registry (JTTR), which details combat injury type and location.

### Data handling

#### Data processing

Data were pre-processed with a bespoke Python workflow. For the nested study addressing the second aim, this workflow was applied exclusively to the Injured cohort excluding those with lower limb loss. Focusing on this sub-cohort allowed assessment of heterogeneity in musculoskeletal outcomes without the confounding influence of limb loss.

#### Cohort curation

Participants labelled (Injured, non-amputee) in the study metadata were retained. Questionnaire variables were kept at total- or domain-score level to minimise multicollinearity and reduce the risk of model over-fitting; individual item scores were discarded. All musculoskeletal-adjacent variables listed in Table 3 were imported and subjected to range and type checks. Records with missing or implausible values in key demographic or musculoskeletal fields (e.g., body mass, New Injury Severity Score) were removed.

#### Modality construction

Features were organised into modality-specific matrices—self-reported outcomes, clinical assessments, and imaging-derived measures—consistent with the sparse GFA framework. Continuous variables were standardised to zero mean and unit variance, and categorical variables were one-hot encoded. Outliers were retained because the hierarchical Bayesian formulation of sparse GFA is robust to extreme observations, and such extremes are clinically informative in trauma cohorts.

To ensure comparable latent-factor estimation across modalities, only participants with complete data for every retained modality were included in the final analysis set. The resulting multimodal matrix constituted the input to the sparse GFA model described in the subsequent section and represents a harmonised, quality-controlled view of musculoskeletal health in the Injured non-amputee group.

### Data analysis

#### Sparse GFA

Sparse GFA identifies latent factors in subgroups that capture the relationships within multimodal data (31). Sparse GFA is an extension of GFA (63) where regularised horseshoe priors are added over the loading matrices and latent variables. The regularised horseshoe prior (64) is a popular shrinkage prior used in sparse Bayesian regression to ensure that small coefficients are heavily shrunk towards zero, while large coefficients are regularised but remain large. In this way, sparse GFA improves model interpretability and identifies latent variables differently expressed across the sample.

In this study, sparse GFA was fitted using a Markov chain Monte Carlo algorithm with five sampling chains and 4,000 samples (the first 1,000 were discarded as warm-up). The inference was randomly initialised five times and the best initialisation was selected to maximise the expected log joint posterior density. All sampling chains were initialised with 20 factors. More details about the experiment setup, are in Ferreira et al (31).

We used the approach proposed by Ferreira et al (31) to select robust factors, i.e., the factors are first averaged over the posterior samples within a sampling chain and then compared to the factors obtained in other sampling chains using cosine similarity. Two factors were considered the same if the cosine similarity between them was greater than 0.80. A factor was considered robust if it had been obtained in at least half of the sampling chains.

#### Interpretation

For each participant, sparse GFA calculates a distribution across the latent variables, showing how a particular participant is represented in each component. For each component, we also obtain a representation across the features in each data modality. This provides a loading matrix showing the importance of each feature in each component for each participant, which we refer to as *data components*. These data components allow interpretation of the results (e.g., a positive [more red] value for a particular participant can be interpreted as scoring high in a task).

In Study 1, data components were considered where there was a clear difference between Injured and Uninjured groups (e.g., Oswestry Disability Index (ODI) scores were largely red for the Injured group and blue for the Uninjured group; Figure 1a). The hypothesis regarding this difference was inherent due to the known way the data was split into groups. The Injured group contained a sub-group of 157 men who sustained injuries resulting in limb loss. They were not labelled but could be identified through limb loss-specific outcomes (e.g., phantom limb pain), or where non-specific outcomes (e.g., low bone mineral density) were shown in combination with limb loss-specific measures.

In Study 2, a nested sub-set of Injured participants without lower limb loss injuries were analysed without an inherent group structure to demonstrate a real-world application of sparse GFA that could potentially generate de novo hypotheses on causation or correlation. Loading matrices are analysed to identify a group structure (e.g., sub-group have poor scores across specific outcomes). Next, patterns in the data are investigated and used to develop a novel hypothesis. In this example, the following steps were taken:

1. Group structure identified sub-group with poor scores across specific outcomes
2. Participants were manually divided into those with and without poor scores for those specific outcomes
3. Injury location, demographic information, and injury severity visually and statistically compared between groups
4. Overall health EQ-5D-5L score visually compared between injury location groups

#### Statistical analysis

JTTR records were unavailable for 41 Injured participants who were subsequently excluded from analysis involving injury location or New Injury Severity Scores (NISS; higher score = greater severity).

In the first study, 1 Uninjured participant was excluded due to non-combat related lower limb loss following matching.

In the second study, where statistical comparisons of continuous variables were made between the groups, data was visually assessed for normality using histograms, and an independent t-test or Mann-Whitney U test was used, as appropriate. Boxplots were compared visually for trends. Descriptive data and statistical analysis were processed using Stata SE version 18.5.

### Code and data availability

The code was adapted from Ferreira et al (31) and is available at https://github.com/ferreirafabio80/sgfa.

Given their unusually sensitive nature, data have not been made widely available. Requests for data will be considered on a case-by-case basis and subject to UK Ministry of Defence clearance via.

## Acknowledgements

We wish to thank all of the research and clinical staff at Headley Court, Stanford Hall and the Defence Medical Services who have helped with the ADVANCE study, including Maria-Benedicta Edwards, Helen Blackman, Melanie Chesnokov, Emma Coady, Sarah Evans, Guy Fraser, Meliha Kaya-Barge, Maija Maskunitty, David Pernet, Helen Prentice, Urszula Pucilowaska, Stefan Sprinchmoller, Owen Walker, Eleanor Miller, Lajli Varsani, Anna Varey, Molly Waldron, Danny Weston, Tass White, Seamus Wilson, Louise Young, Dan Dyball, Col Ian Gibb, Surg Cmd David Gray, and Lt Col Edward Sellon. We would also like to thank the ADVANCE study participants for their generous and ongoing donation of time.

## References

1. Harvey NC, Clegg PD, Dennison EM, Greenhaff P, Griffin SJ, Gregson CL, et al. UKRI MRC National Musculoskeletal Ageing Network: strategic prioritisation to increase healthy lifespan and minimise physical frailty. Arch Osteoporos. 2022;17(1):147.

2. Boer CG, Hatzikotoulas K, Southam L, Stefansdottir L, Zhang Y, Coutinho de Almeida R, et al. Deciphering osteoarthritis genetics across 826,690 individuals from 9 populations. Cell. 2021;184(18):4784–818 e17.

3. Osborne A, Blake C, Fullen BM, Meredith D, Phelan J, McNamara J, et al. Risk factors for musculoskeletal disorders among farm owners and farm workers: a systematic review. Am J Ind Med. 2012;55(4):376–89.

4. Lovalekar M, Hauret K, Roy T, Taylor K, Blacker SD, Newman P, et al. Musculoskeletal injuries in military personnel-Descriptive epidemiology, risk factor identification, and prevention. J Sci Med Sport. 2021;24(10):963–9.

5. Whittaker JL, Losciale JM, Juhl CB, Thorlund JB, Lundberg M, Truong LK, et al. Risk factors for knee osteoarthritis after traumatic knee injury: a systematic review and meta-analysis of randomised controlled trials and cohort studies for the OPTIKNEE Consensus. Br J Sports Med. 2022;56(24):1406–21.

6. Behan F, Bennett A, Schofield S, Miller E, Bull A. Osteoarthritis after major combat trauma: the ADVANCE study. OARSI World Congress of Osteoarthritis; Vienna, Austria: Osteoarthritis & Cartilage; 2024. p. S265.

7. Gwinnutt JM, Wieczorek M, Balanescu A, Bischoff-Ferrari HA, Boonen A, Cavalli G, et al. 2021 EULAR recommendations regarding lifestyle behaviours and work participation to prevent progression of rheumatic and musculoskeletal diseases. Ann Rheum Dis. 2023;82(1):48–56.

8. Dunn M, Rushton AB, Mistry J, Soundy A, Heneghan NR. The biopsychosocial factors associated with development of chronic musculoskeletal pain. An umbrella review and meta-analysis of observational systematic reviews. PLoS One. 2024;19(4):e0294830.

9. Taanila H, Suni JH, Kannus P, Pihlajamaki H, Ruohola JP, Viskari J, et al. Risk factors of acute and overuse musculoskeletal injuries among young conscripts: a population-based cohort study. BMC Musculoskelet Disord. 2015;16:104.

10. van Poppel D, van der Worp M, Slabbekoorn A, van den Heuvel SSP, van Middelkoop M, Koes BW, et al. Risk factors for overuse injuries in short- and long-distance running: A systematic review. J Sport Health Sci. 2021;10(1):14–28.

11. Evans SL, Owen R, Whittaker G, Davis OE, Jones ES, Hardy J, et al. Non-contact lower limb injuries in Rugby Union: A two-year pattern recognition analysis of injury risk factors. PLoS One. 2024;19(10):e0307287.

12. da Costa BR, Vieira ER. Risk factors for work-related musculoskeletal disorders: A systematic review of recent longitudinal studies. Am J Ind Med. 2010;53(3):285–323.

13. Zebis MK, Andersen LL, Brandt M, Myklebust G, Bencke J, Lauridsen HB, et al. Effects of evidence-based prevention training on neuromuscular and biomechanical risk factors for ACL injury in adolescent female athletes: a randomised controlled trial. Br J Sports Med. 2016;50(9):552–7.

14. Schram B, Orr R, Pope R, Canetti E, Knapik J. Risk factors for development of lower limb osteoarthritis in physically demanding occupations: A narrative umbrella review. J Occup Health. 2020;62(1):e12103.

15. Moen BE, Koefoed VF, Bondevik K, Haukenes I. A survey of occupational health in the Royal Norvegian Navy. Int Marit Health. 2008;59(1-4):35–44.

16. Collaborators GBDOMD. Global, regional, and national burden of other musculoskeletal disorders, 1990-2020, and projections to 2050: a systematic analysis of the Global Burden of Disease Study 2021. Lancet Rheumatol. 2023;5(11):e670–e82.

17. Disease GBD, Injury I, Prevalence C. Global, regional, and national incidence, prevalence, and years lived with disability for 354 diseases and injuries for 195 countries and territories, 1990-2017: a systematic analysis for the Global Burden of Disease Study 2017. Lancet. 2018;392(10159):1789–858.

18. Data Versus Arthritis. The state of musculoskeletal health 2024. Arthritis and other musculoskeletal conditions in numbers. 2024.

19. Murray CJ, Vos T, Lozano R, Naghavi M, Flaxman AD, Michaud C, et al. Disability-adjusted life years (DALYs) for 291 diseases and injuries in 21 regions, 1990-2010: a systematic analysis for the Global Burden of Disease Study 2010. Lancet. 2012;380(9859):2197–223.

20. DALYs GBD, Collaborators H. Global, regional, and national disability-adjusted life-years (DALYs) for 359 diseases and injuries and healthy life expectancy (HALE) for 195 countries and territories, 1990-2017: a systematic analysis for the Global Burden of Disease Study 2017. Lancet. 2018;392(10159):1859–922.

21. Hinton G. Deep Learning-A Technology With the Potential to Transform Health Care. JAMA. 2018;320(11):1101–2.

22. Rajkomar A, Dean J, Kohane I. Machine Learning in Medicine. N Engl J Med. 2019;380(14):1347–58.

23. Topol EJ. High-performance medicine: the convergence of human and artificial intelligence. Nat Med. 2019;25(1):44–56.

24. Ipp E, Liljenquist D, Bode B, Shah VN, Silverstein S, Regillo CD, et al. Pivotal Evaluation of an Artificial Intelligence System for Autonomous Detection of Referrable and Vision-Threatening Diabetic Retinopathy. JAMA Netw Open. 2021;4(11):e2134254.

25. Ardila D, Kiraly AP, Bharadwaj S, Choi B, Reicher JJ, Peng L, et al. End-to-end lung cancer screening with three-dimensional deep learning on low-dose chest computed tomography. Nat Med. 2019;25(6):954–61.

26. McKinney SM, Sieniek M, Godbole V, Godwin J, Antropova N, Ashrafian H, et al. International evaluation of an AI system for breast cancer screening. Nature. 2020;577(7788):89–94.

27. Komorowski M, Celi LA, Badawi O, Gordon AC, Faisal AA. The Artificial Intelligence Clinician learns optimal treatment strategies for sepsis in intensive care. Nat Med. 2018;24(11):1716–20.

28. Kadirvelu B, Gavriel C, Nageshwaran S, Chan JPK, Nethisinghe S, Athanasopoulos S, et al. A wearable motion capture suit and machine learning predict disease progression in Friedreich’s ataxia. Nat Med. 2023;29(1):86–94.

29. Ricotti V, Kadirvelu B, Selby V, Festenstein R, Mercuri E, Voit T, et al. Wearable full-body motion tracking of activities of daily living predicts disease trajectory in Duchenne muscular dystrophy. Nat Med. 2023;29(1):95–103.

30. Greener JG, Kandathil SM, Moffat L, Jones DT. A guide to machine learning for biologists. Nat Rev Mol Cell Biol. 2022;23(1):40–55.

31. Identifying latent disease factors differently expressed in patient subgroups using group factor analysis [Internet]. arxiv. 2024 [cited 21/10/2024]. Available from: https://arxiv.org/abs/2410.07890.

32. Bennett AN, Dyball DM, Boos CJ, Fear NT, Schofield S, Bull AMJ, et al. Study protocol for a prospective, longitudinal cohort study investigating the medical and psychosocial outcomes of UK combat casualties from the Afghanistan war: the ADVANCE Study. BMJ Open. 2020;10(10):e037850.

33. McMenemy L, Behan FP, Kaufmann J, Cain D, Bennett AN, Boos CJ, et al. Association Between Combat-Related Traumatic Injury and Skeletal Health: Bone Mineral Density Loss Is Localized and Correlates With Altered Loading in Amputees: the Armed Services Trauma Rehabilitation Outcome (ADVANCE) Study. J Bone Miner Res. 2023.

34. Watson FCE, Kedgley AE, Schofield S, Behan FP, Boos CJ, Fear NT, et al. Upper Limb Function in People With Upper and Lower Limb Loss 8 Years Postinjury: The Armed Services Trauma Outcome Study (ADVANCE) Cohort Study. Phys Ther. 2024.

35. O’Sullivan O, Stocks J, Schofield S, Bilzon J, Boos CJ, Bull AMJ, et al. Association of serum biomarkers with radiographic knee osteoarthritis, knee pain and function in a young, male, trauma-exposed population - Findings from the ADVANCE study. Osteoarthritis Cartilage. 2024.

36. Vollert J, Kumar A, Coady EC, Cullinan P, Dyball D, Fear NT, et al. Pain after combat injury in male UK military personnel deployed to Afghanistan. Br J Anaesth. 2024;132(6):1285–92.

37. Watson FCE, A NB, McGregor A, Behan F, N TF, C JB, et al. Biopsychosocial factors and low back pain in military personnel with lower limb loss: the ADVANCE study. BMJ Mil Health. 2025.

38. Watson FCE, Agricola R, Bennett A, Bull A. Prevalence, and 3-year progression and incidence of radiographic hip osteoarthritis in a yuong military population post major trauma: the ADVANCE study. Osteoarthritis & Cartilage. 2025;33:S56–S7.

39. Bluher M. Obesity: global epidemiology and pathogenesis. Nat Rev Endocrinol. 2019;15(5):288–98.

40. Warden D. Military TBI during the Iraq and Afghanistan wars. J Head Trauma Rehabil. 2006;21(5):398–402.

41. McPherson AL, Nagai T, Webster KE, Hewett TE. Musculoskeletal Injury Risk After Sport-Related Concussion: A Systematic Review and Meta-analysis. Am J Sports Med. 2019;47(7):1754–62.

42. Reneker JC, Babl R, Flowers MM. History of concussion and risk of subsequent injury in athletes and service members: A systematic review and meta-analysis. Musculoskelet Sci Pract. 2019;42:173–85.

43. Roach MH, Aderman MJ, Ross JD, Kelly TF, Malvasi SR, Posner MA, et al. Risk of Upper Extremity Musculoskeletal Injury Within the First Year After a Concussion. Orthop J Sports Med. 2023;11(5):23259671231163570.

44. Brown S, Hawker G, Beaton D, Colantonio A. Long-term musculoskeletal complaints after traumatic brain injury. Brain Inj. 2011;25(5):453–61.

45. Shumski EJ, Oh J, Schmidt JD, Lynall RC. Trunk and Lower Extremity Biomechanics in Female Athletes With and Without a Concussion History. J Athl Train. 2024;59(7):751–61.

46. Shumski EJ, Schmidt JD, Lynall RC. Cognition Uniquely Influences Dual-Task Tandem Gait Performance Among Athletes With a Concussion History. Sports Health. 2024;16(4):542–50.

47. Nawi MAA, Noor NFM, Shaari R, Khaleel AK, Lazin MAM, Sulaiman IM, et al. The patterns of facial fractures in traumatic brain injury (TBI) patients using ordinal regression: a retrospective study of five years. AIMS Neurosci. 2022;9(3):345–57.

48. Feng YS, Kohlmann T, Janssen MF, Buchholz I. Psychometric properties of the EQ-5D-5L: a systematic review of the literature. Qual Life Res. 2021;30(3):647–73.

49. Defence Mo. Freedom of Information request 2015/07390. In: Statistics D, editor. 2015.

50. Colangelo PM, Welch DW, Rich DS, Jeffrey LP. Two methods for estimating body surface area in adult amputees. Am J Hosp Pharm. 1984;41(12):2650–5.

51. Haycock GB, Schwartz GJ, Wisotsky DH. Geometric method for measuring body surface area: a height-weight formula validated in infants, children, and adults. J Pediatr. 1978;93(1):62–6.

52. Kellgren JH, Lawrence JS. Radiological assessment of osteo-arthrosis. Ann Rheum Dis. 1957;16(4):494–502.

53. Altman RD, Gold GE. Atlas of individual radiographic features in osteoarthritis, revised. Osteoarthritis Cartilage. 2007;15 Suppl A:A1–56.

54. Gailey RS, Roach KE, Applegate EB, Cho B, Cunniffe B, Licht S, et al. The amputee mobility predictor: an instrument to assess determinants of the lower-limb amputee’s ability to ambulate. Arch Phys Med Rehabil. 2002;83(5):613–27.

55. Poddubnyy D, van Tubergen A, Landewe R, Sieper J, van der Heijde D, Assessment of SpondyloArthritis international S. Development of an ASAS-endorsed recommendation for the early referral of patients with a suspicion of axial spondyloarthritis. Ann Rheum Dis. 2015;74(8):1483–7.

56. Herdman M, Gudex C, Lloyd A, Janssen M, Kind P, Parkin D, et al. Development and preliminary testing of the new five-level version of EQ-5D (EQ-5D-5L). Qual Life Res. 2011;20(10):1727–36.

57. Fairbank JC, Couper J, Davies JB, O’Brien JP. The Oswestry low back pain disability questionnaire. Physiotherapy. 1980;66(8):271–3.

58. Roos EM, Lohmander LS. The Knee injury and Osteoarthritis Outcome Score (KOOS): from joint injury to osteoarthritis. Health Qual Life Outcomes. 2003;1:64.

59. Germann G, Wind G, Harth A. The DASH (Disability of Arm-Shoulder-Hand) Questionnaire - a new instrument for evaluating upper extremity treatment outcome. Handchir Mikrochir Plast Chir. 1999;31(3):149–52.

60. Craig CL, Marshall AL, Sjostrom M, Bauman AE, Booth ML, Ainsworth BE, et al. International physical activity questionnaire: 12-country reliability and validity. Med Sci Sports Exerc. 2003;35(8):1381–95.

61. Ryall NH, Eyres SB, Neumann VC, Bhakta BB, Tennant A. The SIGAM mobility grades: a new population-specific measure for lower limb amputees. Disabil Rehabil. 2003;25(15):833–44.

62. Hanspal RS, Fisher K, Nieveen R. Prosthetic socket fit comfort score. Disabil Rehabil. 2003;25(22):1278–80.

63. Virtanen S, Klami A, Khan S, Kaski S. Bayesian Group Factor Analysis. In: Lawrence ND, Girolami M, editors. Proceedings of Machine Learning Research2012. p. 1269–77.

64. Piironen J, Vehtari A. Sparsity information and regularization in the horseshoe and other shrinkage priors. Electronic Journal of Statistics. 2017;11(2). 27

